# Brain health across the entire glycaemic spectrum: the UK Biobank

**DOI:** 10.1101/2020.02.18.20024471

**Authors:** Victoria Garfield, Aliki-Eleni Farmaki, Sophie V. Eastwood, Rohini Mathur, Christopher T. Rentsch, Krishnan Bhaskaran, Liam Smeeth, Nish Chaturvedi

## Abstract

**Objective:** To understand the relationship across the glycaemic spectrum, with brain health. We hypothesised that individuals with increasingly higher HbA_1c_ would be more likely to have worse brain health outcomes in comparison to normoglycaemic individuals.

**Methods:** UK Biobank participants. HbA_1c_ and diabetes diagnosis define baseline glycaemic categories. Outcomes: incident vascular dementia (VD), Alzheimer’s dementia (AD), hippocampal volume (HV), white matter hyperintensity (WMH) volume, cognitive function and decline. Reference group: normoglycaemic individuals (HbA_1c_ 35-<42 mmol/mol).

**Results:** Pre- and known diabetes increased incident VD, (HR 1.54, 95%CI=1.04;2.28 and 2.97, 95%CI=2.26;3.90). Known diabetes increased AD risk (HR 1.84, 95%CI=1.44;2.36). Pre- and known diabetes elevated risks of cognitive decline (OR 1.42, 1.48;2.96 and 1.39, 1.04;1.75). Pre-diabetes, undiagnosed and known diabetes conferred higher WMH volumes (4%, 26%, 5%,) and lower HV (22.4mm^3^, 15.2mm^3^, 62.2mm^3^). Low-normal HbA_1c_ had 2% lower WMH volume and 13.6mm^3^ greater HV.

**Interpretation:** Pre and known diabetes increase VD risks; known diabetes increases AD risk. Low-normal HbA_1c_ associates with more favourable neuroimaging outcomes. Our findings may have implications for cardiovascular medication in hyperglycaemia for brain health.

## INTRODUCTION

Type-2 diabetes and, more generally, hyperglycaemic states, have been associated with poorer cognitive function (such as learning and memory)^1,2^, increased risk of dementia^2,3^ and alterations in key brain structures, particularly the hippocampus^4^. It is also important to explore how low-normal levels (vs. normal glycaemic levels) of glycated haemoglobin (HbA_1c_) relate to brain health outcomes, which has not been investigated in a population-based study, to date. A previous paper explored the cross sectional association between baseline diabetes and two cognition measures in the UK Biobank (reaction time and visual memory)^5^. The authors found that diabetes was associated with poorer scores on the reaction time test, but paradoxically, better scores on the visual memory test. They did not explore other brain health outcomes or lesser glycaemic states.

Memory loss is the most conclusively reported adverse effect of hyperglycaemia on cognitive function^6^, yet hyperglycaemia also associates with worse processing speed, attention, concentration and executive functions ^7^. Hippocampal atrophy is a crucial feature of age-related memory loss and the hippocampus is reportedly more vulnerable to the neurotoxic consequences of diabetes^8,9^. Evidence relating diabetes to the presence and progression of white matter hyperintensities is equivocal^10^, but some research suggests that those with diabetes have greater volumes of white matter hyperintensities^11,12^. Although there have been numerous studies in this area, the role of glycaemia in brain health across the entire glycaemic spectrum remains unclear, in particular no studies have investigated how lesser hyperglycaemic states relate to these outcomes, as most studies have focused on diagnosed diabetes only. Thus, our aim was to investigate, in a single large-scale study, the associations between five glycaemic states across the entire spectrum (low-normal HbA_1c_, normoglycaemia, pre-diabetes, undiagnosed diabetes and known diabetes) and a breadth of brain health outcomes including: Alzheimer’s dementia (AD) risk, vascular dementia (VD) risk, baseline cognitive function and cognitive decline, hippocampal volume, and white matter hyperintensities volume in the UK Biobank. We hypothesised that those with increasingly higher HbA_1c_ would have worse outcomes compared to those with normal glycaemic levels.

## METHODS

### Sample

Full details of the UK Biobank (UKB) cohort have been described elsewhere^13^. Briefly, UKB consists of ∼500,000 men and women from the general UK population between 2006-2010, aged between 40 and 69 years of age at baseline. Participants have detailed phenotype data on physical (subjective and objective), mental and lifestyle measures, as well as linkages to routinely collected data (e.g. deaths, hospital admissions and cancer registers). Figure 1 depicts our study design.

**Fig 1.**
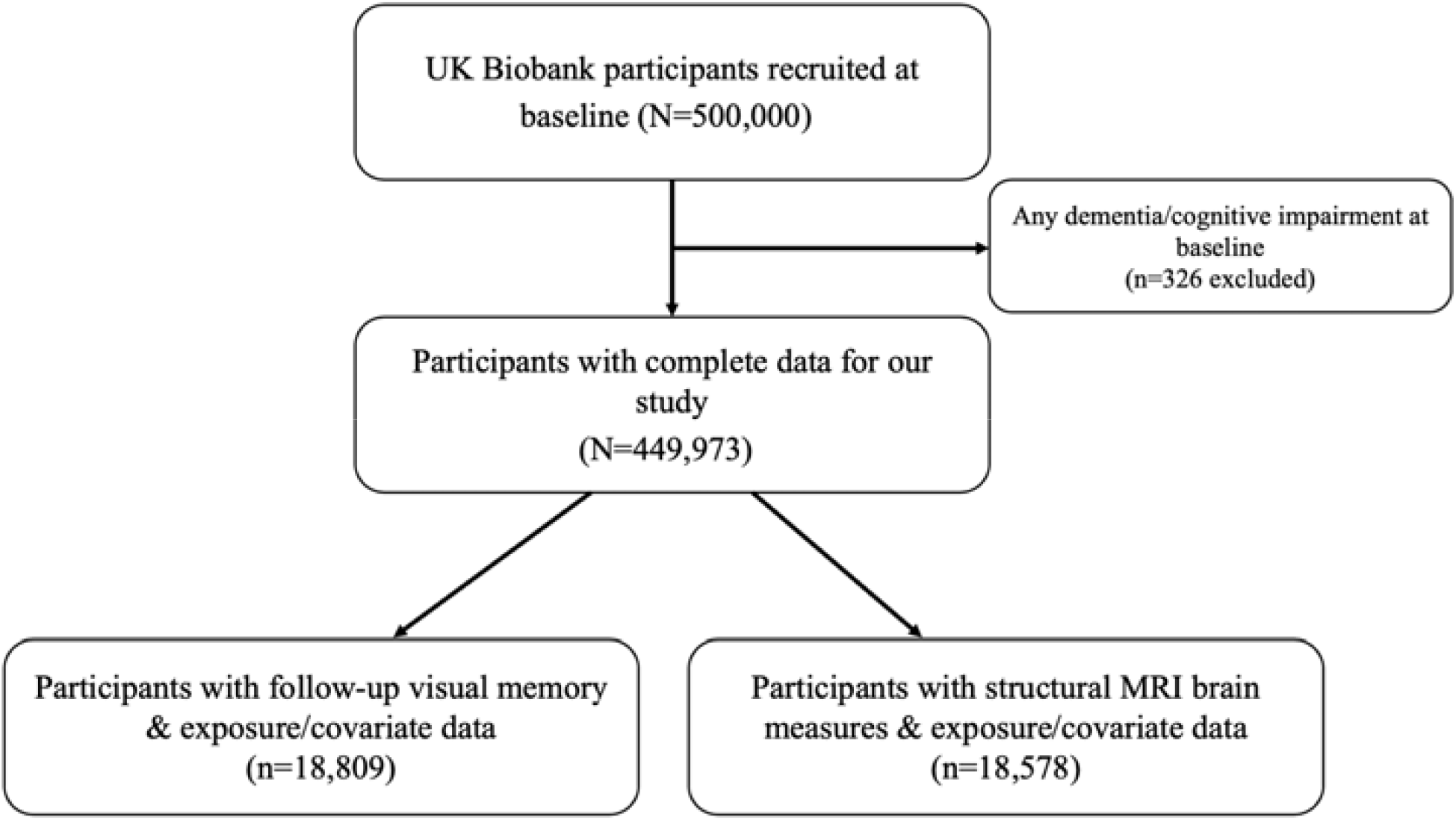
Study design.

### Informed consent and ethical approval

UK Biobank received ethical approval from the North West Multi-centre Research Ethics Committee (MREC) and informed consent has been obtained from participants.

### Type-2 diabetes mellitus (diabetes)

Exposure status was defined using baseline data on diabetes and HbA_1c_. Diabetes was defined using an algorithm of self-report doctor diagnosis and/or medication; this algorithm has been validated against primary care data^14^ HbA_1c_ assays were performed using five Bio-Rad Variant II Turbo analysers, outlined in detail in the UK Biobank protocol^15^. In this study, values greater than 200 mmol/mol were excluded (n=5), as they were considered to be outliers and clinically implausible. For our analyses we divided participants into the following categories: known diabetes, undiagnosed diabetes (≥48 mmol/mol), pre-diabetes (42-<48 mmol/mol), normoglycaemic (≥35 & <42 mmol/mol), and low HbA_1c_ (<35 mmol/mol) – based on criteria by Ginde and colleagues ^16^.

### Dementia

Dementia at baseline was captured using ICD-10 codes in linked hospital episode statistics (HES) data. Incident dementia was algorithmically defined with the method described in Wilkinson et al. ^17^, which was based on linked UK hospital admission, mortality and primary care data. Here we focus on vascular dementia (n=412) and AD (n=749); frontotemporal dementia cases were excluded (n=95).

### Structural brain MRI outcomes

Structural brain MRI scans have been performed in a subsample of UKB participants using standard protocols, details of which are published^18^. Post-processed measures (provided by UKB) used in this study included: hippocampal volume (mm^3^), normalised for head size and total volume (mm^3^) of white matter hyperintensities (WMH, mm^3^). WMH volume was log-transformed as it was positively skewed; thus, we report exponentiated betas for this outcome to ease interpretation. The total maximum sample size for participants with these outcomes in our study was n=18,578.

### Cognitive function

For cognitive function we pragmatically selected two measures with adequate sample sizes to represent distinct cognitive domains, namely reaction time (RT) and visual memory. In the visual memory test, respondents had to identify matches from six pairs of cards after memorising their positions on the screen. The number of incorrect matches (errors made) was then recorded, whereby a higher number indicates poorer visual memory. RT was measured as the mean time (in milliseconds) taken to correctly identify matches from 12 rounds of the card game ‘Snap’, where a longer time indicates slower RTs. As per Lyall et al., (2016)^19^ reaction time (RT) was transformed using a log transformation (ln) and visual memory was transformed using an ln+1 equation (due to zero-value inflation). The total sample size for the reaction time and visual memory baseline analyses was 449,973.

### Cognitive decline

Using data from a subset of participants with both baseline and follow-up measures of cognitive function, cognitive decline was determined using the Standardised Regression Based method^20^. This included regressing follow-up visual memory on baseline visual memory, age, sex, years of education, and time between the two assessments. Subsequently, those whose standardised residual was greater than (absolute value) 1.96 (0.05 type-1 error rate) were assigned as having cognitive decline. Only a proportion of the UKB participants had follow-up visual memory data and complete covariate data (n=18,809).

### Covariates

Demographics such as age (years), sex, ethnicity (White European, Asian/Asian British, Black/Black British, Other), deprivation (quintiles of Townsend deprivation index, from ‘least deprived’ to ‘most deprived’), and educational attainment (derived as years of full-time education completed, as per qualifications based on coding from the International Standard Classification of Education ^21^) were included. Health behaviours included smoking status (never, current smoker and ex-smoker). Health measures included body mass index (BMI) in kg/m^2^, baseline cardiovascular disease (CVD – assigned using baseline self-report, nurse interview and linked hospital inpatient data between 2006 and 2010), anti-hypertensive medication and statin use; medications were captured and classified according to British National Formulary (BNF) chapters.

### Exclusion criteria

Those who had AD, vascular or frontotemporal dementia or cognitive impairment prior to their recorded date of baseline assessment (2006-2010), as captured by self-report, nurse interview or HES were excluded.

### Missing data

There were missing data across several variables, all of which had <10% missingness and for this reason we used complete case analysis for this study. The missing data were as follows: ethnicity n=2275, BMI n=3260, reaction time n=5776, visual memory n=4627, deprivation n=623, smoking n=1918, HbA_1c_ n=34,594, antihypertensives and statins n=8589, educational attainment n=9133.

### Statistical analyses

Analyses were performed in RStudio, version 1.1.456 and STATA version 15.

### Modelling approach

#### Cross-sectional cognitive function and structural brain analyses

In the cross-sectional analyses, glycaemia was entered as an exposure and four linear regressions were fitted to explore the relationship with baseline cognition outcomes (reaction time and visual memory). Model 1 consisted of adjustment for demographic measures (age + sex + deprivation + educational attainment + ethnicity), Model 2 was additionally adjusted for standard cardiovascular risk factors (smoking + BMI + CVD + anti-hypertensives + statins). Our modelling approach was identical for structural brain outcomes (hippocampal volume and volume of WMH).

#### Cognitive decline analyses

Only 4% of UKB participants underwent follow-up cognition testing, so our analyses of cognitive decline were restricted to this sub-population. The same models were fitted as in the analyses of baseline data. Logistic regression was used to investigate the association between glycaemia and binary cognitive decline, with the same modelling strategy as above.

#### Dementia analyses

Cox proportional hazards models were used to examine the relationships between glycaemia and a) AD and b) vascular dementia. The time scale was time since study entry and participants were followed up until 31 March 2017. The same modelling strategy was used, as described above. The proportional hazards assumption was assessed using the global test to evaluate the interaction of each covariate with time, alongside Schoenfeld residuals.

## RESULTS

### Sample characteristics

449,973 individuals were included in the study, of whom 210,309 had low-normal HbA_1c_ levels, 198,969 had normoglycaemic levels, 15,229 had pre-diabetes, 3279 had undiagnosed diabetes and 22,187 had known diabetes. Those with prediabetes and known diabetes were older than the other groups. Those with diabetes (undiagnosed and known) were more likely to be ex-smokers, reside in the most deprived quintile and have higher BMIs (Table 1). Those with known diabetes were most likely to be taking antihypertensives and statins at baseline and had the highest prevalence of CVD.

**Table 1.**
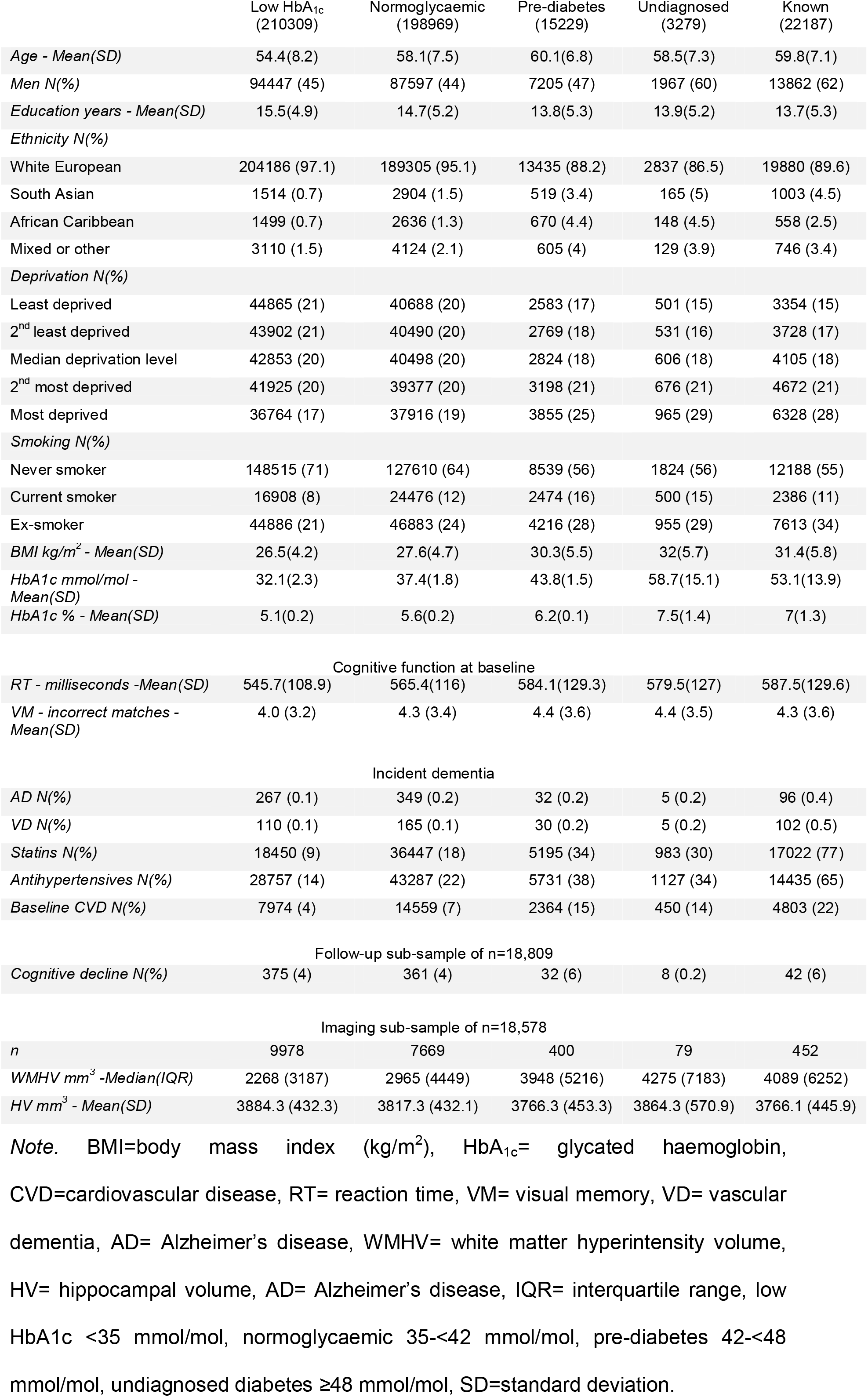
Baseline characteristics and outcomes across the glycaemic spectrum, N= 449,973

### Glycaemia and AD, and vascular dementia (VD)

We do not present results from the undiagnosed diabetes group, as the number of cases for both AD and VD was <20. Pre-diabetes and low-normal HbA_1c_ were not associated with AD in a basic or fully-adjusted model (Fig 2). However, known diabetes was strongly associated with excess AD risk on minimal adjustment and this remained robust in a fully-adjusted model (HR 1.84, 95%CI=1.44;2.36). People with pre-diabetes had elevated risks of VD, as did those with known diabetes (HR 1.75, 95% CI=1.19;2.59 and HR 3.73, 95% CI=2.90;4.80, respectively) (Fig 2), but low-normal HbA_1c_ was not associated with VD (Fig 2).

**Fig. 2.**
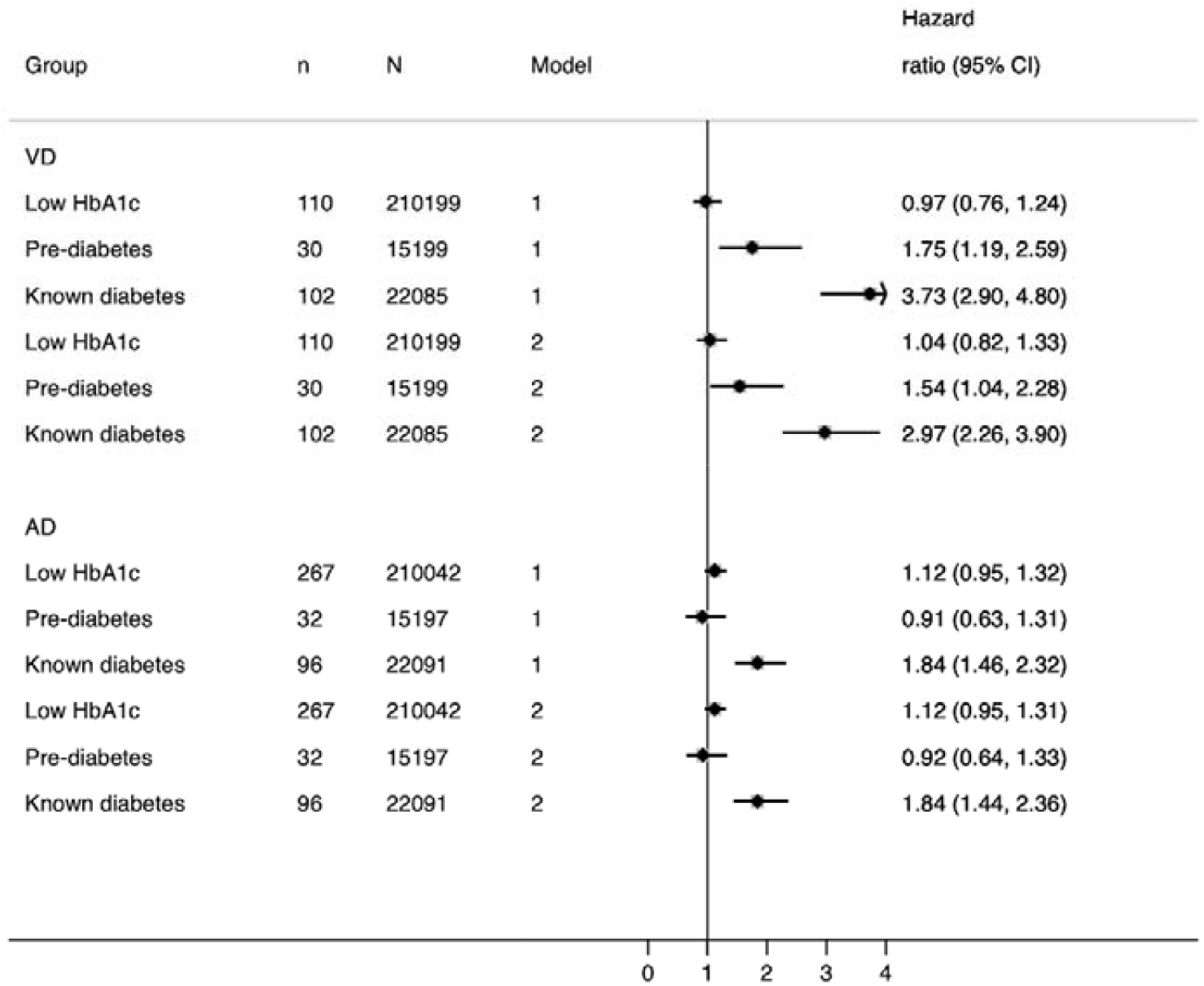
Association between glycaemia and incident Alzheimer’s and vascular dementia in UK Biobank (N=449,973) *Note*. Model 1= adjusted for age + sex + deprivation + ethnicity + educational attainment, Model 2= Model 1 + BMI + CVD + statins + antihypertensives + smoking, 95%CI = 95% confidence interval.

Adjustment for health-related measures attenuated associations between glycaemia and VD, but this remained large at 54% increased risk of VD in pre-diabetes and almost 3-fold excess risk in known diabetes. The key factor responsible for accounting for excess risk for both pre-diabetes and known diabetes in multi-variate models was antihypertensive therapy. Model 1 HRs were 1.75 (95% CI=1.19;2.59) and 3.73 (95% CI= 2.90;4.80) for pre-diabetes and known diabetes, respectively. Additional adjustment for antihypertensive therapy only (in addition to Model 1), resulted in HR 1.61 (95% CI= 1.09; 2.39) for pre-diabetes and 3.04 (95% CI= 2.34;3.95) for known diabetes. We also performed sensitivity analyses for both AD and VD in which we included both systolic blood pressure (SBP) alongside antihypertensives in our multiply-adjusted models. However, as the results remained qualitatively the same, albeit with less precision due to a smaller number of cases, we do not present these estimates.

### Glycaemia and hippocampal and white matter hyperintensity volumes

Low-normal HbA_1c_ was associated with lower WMH volume, and greater hippocampal volume than normoglycaemic individuals, while pre, undiagnosed and known diabetes were associated with higher WMH volume and lower hippocampal volume (Fig 3). Multivariable adjustment, specifically when antihypertensive therapy was added, markedly attenuated associations for pre and known diabetes, but less so for undiagnosed diabetes, for WMH volume only. Thus pre-diabetes, undiagnosed diabetes and known diabetes were associated with greater WMH volumes (4%, 26% and 5% respectively), and smaller hippocampal volumes (22mm^3^, 15mm^3^, 62mm^3^) in fully-adjusted models. Those with low-normal HbA_1c_ had 2% lower WMH volume, and 14mm^3^ larger hippocampal volumes than normoglycaemic individuals.

**Fig. 3.**
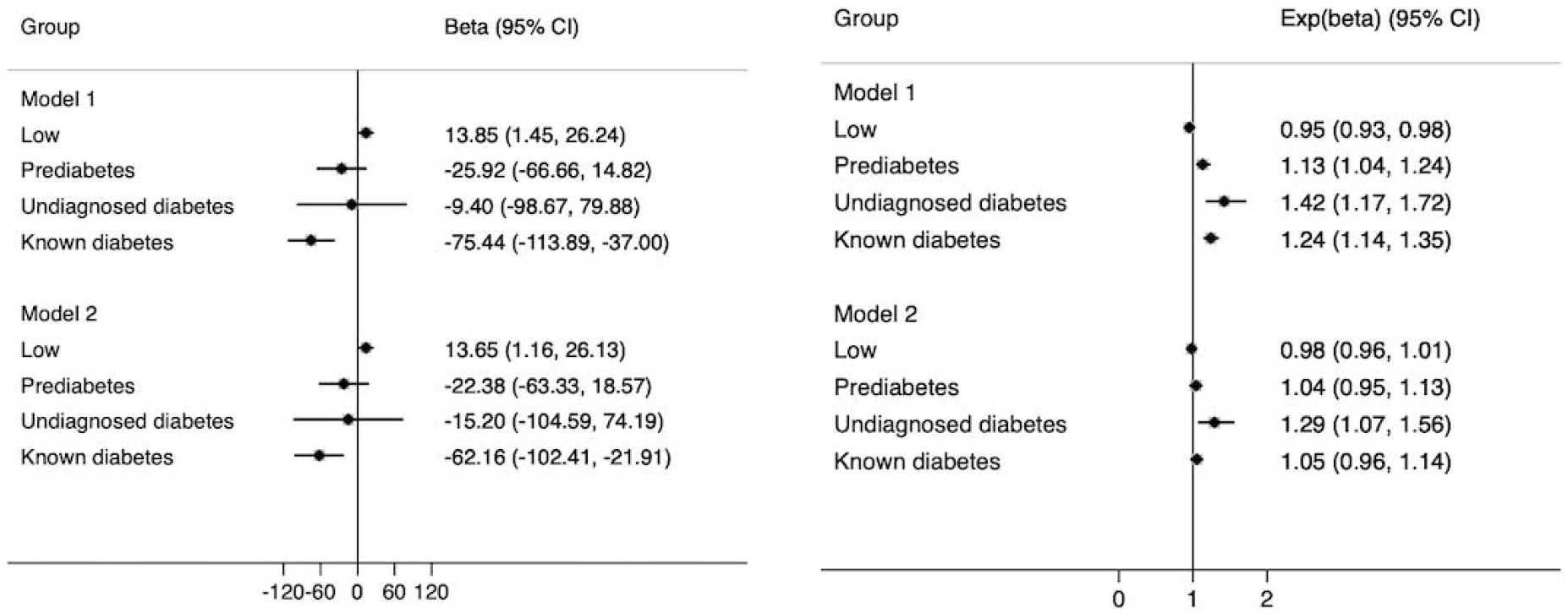
Association between glycaemia and, hippocampal and white matter hyperintensity volumes in a UKB subsample (N=18,578) *Note*. Model 1= adjusted for age + sex + deprivation + ethnicity + educational attainment, Model 2= Model 1 + BMI + CVD + statins + antihypertensives + smoking, 95%CI = 95% confidence interval.

### Glycaemia, baseline reaction time and visual memory, and cognitive decline

Those with low-normal HbA_1c_ had reaction times that were no different to the normoglycaemic group; however, both undiagnosed and known diabetes were associated with a 2% slower reaction time, while pre-diabetes was related to 1% slower reaction times on multivariate adjustment (Table 2). Low-normal HbA_1c_ and undiagnosed diabetes were not associated with visual memory scores, but those with known diabetes made 3% fewer errors, compared to the normoglycaemic group (Table 2). In Model 1 (demographics) pre-diabetes and known diabetes were associated with somewhat greater risk of cognitive decline (Fig 4), but the 95% confidence intervals around the odds ratios were wide. However, in the fully-adjusted model these associations became more pronounced and pre-diabetes and known diabetes related to a 42% and 39% increased risk of cognitive decline, respectively. Upon close inspection of the model, we observed a strong relationship between BMI and cognitive decline, which suggested that those with a higher BMI were less likely to suffer from cognitive decline, OR 0.97 (95%CI = 0.95; 0.99). This remained identical upon multivariate adjustment for age, sex, deprivation, smoking, statins, antihypertensives and CVD.

**Table 2.** Association between glycaemia and baseline cognitive function, N=449,973

| Group | Reaction time | Visual memory |
| --- | --- | --- |
| | Expβ (95% CI) | Expβ (95% CI) |
| Model 1 |  |  |
| Low HbA<sub>1c</sub> | 1.00 (0.99;1.00) | 1.00 (1.00;1.00) |
| Prediabetes | 1.01 (1.01;1.01) | 0.99 (0.98;1.00) |
| Undiagnosed T2DM | 1.01 (1.01;1.02) | 0.99 (0.96;1.01) |
| Known T2DM | 1.02 (1.01;1.02) | 0.97 (0.96;0.98) |
| Model 2 |  |  |
| Low HbA<sub>1c</sub> | 1.00 (0.99;1.00) | 1.00 (0.99;1.00) |
| Prediabetes | 1.01 (1.01;1.01) | 1.00 (0.98;1.01) |
| Undiagnosed T2DM | 1.02 (1.01;1.02) | 1.00 (0.98;1.03) |
| Known T2DM | 1.02 (1.01;1.02) | 0.97 (0.96;0.98) |

*Note.* Model 1= adjusted for age + sex + deprivation + ethnicity + educational
Exp( $\beta$ ) =exponentiated beta, 95% CI = 95% confidence interval.

**Fig. 4.**
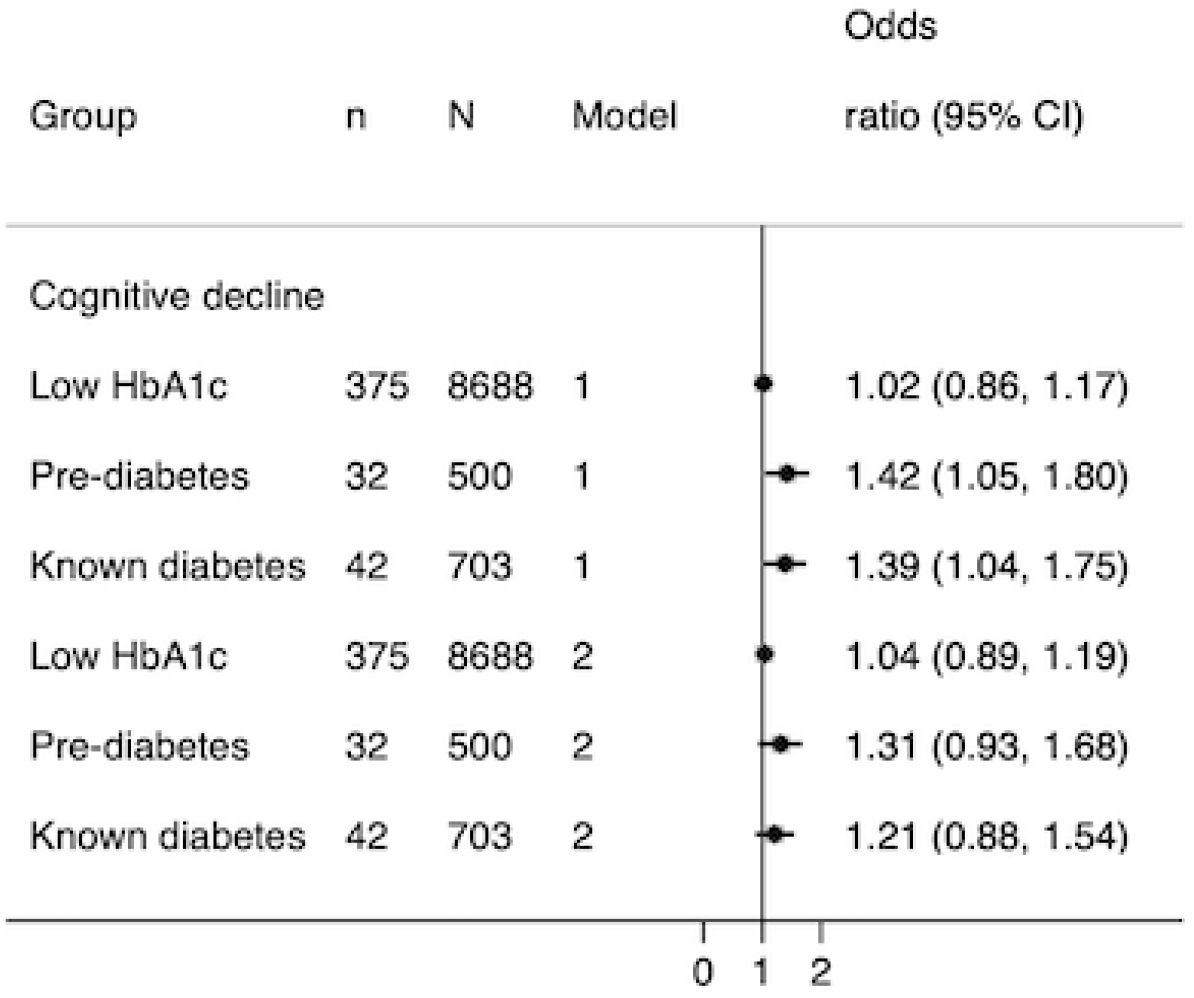
Association between glycaemia and cognitive decline in a UKB subsample (N=18,809) *Note*. Model 1= adjusted for age + sex + deprivation + ethnicity + educational attainment, Model 2= Model 1 + BMI + CVD + statins + antihypertensives + smoking, 95%CI = 95% confidence interval.

## DISCUSSION

In this large sample of middle aged adults we report four key findings; first, people with pre-diabetes and known diabetes have excess risks of clinically important outcomes (cognitive decline and dementia), second, that a key determinant of the excess risk of vascular dementia in association with hyperglycaemia is antihypertensive medication, third, associations between hyperglycaemia and dementia are stronger for vascular than Alzheimer’s dementia and finally, that low-normal levels of glycaemia may be somewhat beneficial in relation to subclinical measures of brain health, such as certain neuroimaging parameters.

It is striking that pre-diabetes and known diabetes both increase the risk of VD, cognitive decline and to a slightly lesser extent AD, in comparison to normoglycaemic individuals. A recent meta-analysis suggests excess dementia risk in pre-diabetes ^22^ but most studies do not make a direct comparison to people with established diabetes, and have been restricted by small numbers of events. Risks of cognitive decline have been more extensively studied, with the majority identifying pre-diabetes as a high-risk state, though few suggest that risks are close to established diabetes ^23,24^. This has important implications for intervention. With greater numbers of individuals surviving to older age, avoidance or at least postponement of dementia is an increasing therapeutic concern, and, much like the finding of excess CVD risks in people with pre-diabetes ^25,26^, prompts consideration of identification and early intervention in such individuals.

Mid-life hypertension increases dementia risk ^27,28^ and is associated with greater WMH volumes ^29^. A recent review of antihypertensive therapy and cerebral small vessel disease (SVD) trials showed that antihypertensive therapy protects against progression of white matter hyperintensities ^30^. That we show attenuation of risk of both VD and WMH volume on adjustment for the greater use of antihypertensive medication in hyperglycaemic states can superficially be interpreted as treatment having adverse, not beneficial, effects. However, we suggest that, in this context, receipt of antihypertensive medication is acting as an indicator of longstanding untreated elevated blood pressure and that therefore, treatment is being instituted too late. This is supported by a recent study which suggests that treatment for hypertension should begin as early as the third decade to potentially reduce risk of disease and early mortality ^31^. Early adulthood blood pressure, measured at around age 43 years, is also more strongly related to WMH volumes at age 70 than blood pressure measured throughout middle age, or indeed contemporaneous with WMH volume assessment ^32^, highlighting the importance of elevated blood pressure even before middle age. The role of even modest elevations in blood pressure, blood pressure trajectories from young adulthood, and early blood pressure lowering intervention, requires exploration in the context of reducing risks of brain pathology.

We show associations between hyperglycaemic states, from pre-diabetes to established diabetes and all of our outcomes, but these appear somewhat stronger with VD and WMH volume than AD and hippocampal volume, as the latter were resistant to adjustment for CVD risk factors. This is in line with evidence that diabetes is associated with greater WMH volume ^11,12^ and a study in the Genetics of Diabetes Audit and Research in Tayside Scotland (GoDARTS) case-control sample, which revealed that those with diabetes had more than a two-fold excess risk of VD, but there was no association with AD ^33^. However, discrimination between VD and AD remains challenging and there have been no studies to date, that have investigated the associations lesser hyperglycaemic states and VD/AD in a single study. That we observed a stronger association between glycaemia and VD and WMH, as opposed to hippocampal volume and AD is perhaps suggestive of two distinct, yet related neurological and vascular pathways. This is in turn is supportive of a ‘two-hit hypothesis’, which has gained popularity more recently ^34^. Briefly, a combination of genetic, environmental and vascular risk factors results in neurovascular dysfunction, alongside damage to arterioles, small arteries and brain capillaries, either through pathways independent of amyloid-ß (hit one) and/or pathways dependent on amyloid-ß (hit two). These pathways converge on blood vessels and can synchronously or independently cause the neuronal dysfunction associated with dementia ^34^. Just how these pathways act synergistically or independently remains unclear.

Another novel finding is that low-normal HbA_1c_ levels were associated with greater hippocampal volume and lower WMH volume, in comparison with normoglycaemic individuals. Participants with low-normal HbA_1c_ tended to be younger and healthier than the other groups, were less likely to be smokers, less likely to reside in higher quintiles of deprivation, had lower prevalence of baseline CVD and fewer of them were on statins or antihypertensives. Adjustment for these factors somewhat attenuated the relationship between low-normal HbA_1c_ and white matter hyperintensity volumes, but this was not the case for hippocampal volume. This may, once again, suggest that distinct mediators operate in the association between glycaemia and AD and atrophy of the brain, compared to factors that mediate the relationship between glycaemia and vascular brain damage. Although our findings preclude us from any temporal or causal claims about this association, it is possible that in middle-aged adults without diabetes (∼54 years) HbA_1c_ levels below 35 mmol/mol could confer some protection against hippocampal atrophy, as well as the presence of white matter hyperintensities. Our findings also indicate that pathways to brain health in association with persistently lower HbA_1c_ in people without diabetes are likely different to those with bouts of hypoglycaemia in people with diabetes. We also observed that pre-diabetes associates with 1% slower reaction times, whereas undiagnosed and known diabetes were associated with 2% slower reaction times. Our finding is supported by an early study of diabetes patients who performed slower on a reaction time task, in comparison to age-matched controls ^35^. We show that there are apparent effects at least cross-sectionally, in pre-diabetes and undiagnosed diabetes, in comparison to normoglycaemia. The association we observed between glycaemia and visual memory was somewhat paradoxical, as known diabetes was associated with 3% fewer incorrect matches on this task. It is possible, however, that other factors common to individuals with diabetes (e.g. effects of medication to control glycaemia) could confer some protection against poorer visual memory.

In the 18,809 participants who had follow-up visual memory data we found that pre-diabetes and known diabetes conferred a 42% and 39% excess risk of cognitive decline on multivariate adjustment. While only 32 people with pre-diabetes, and 42 people with known diabetes experienced cognitive decline during the study follow-up, the fact that both hyperglycaemic states were associated with adverse effects on brain health compared to normoglycaemic individuals adds confidence to our conclusion that hyperglycaemia adversely effects cognitive function, in line with previous observations ^36^. We observed that adjustment for BMI substantially increased the odds ratios from our demographics-only model, such that individuals with a higher BMI were less likely to suffer from cognitive decline. This may relate to the ‘obesity paradox’, whereby those with higher BMIs have lower mortality rates than normal weight individuals, for which several explanations have been proposed^36^.

Our study possesses some important strengths. UK Biobank is one of the largest studies to have data on HbA_1c_ across the entire glycaemic spectrum, cognitive function, dementia sub-types and structural brain MRI measures. We used validated algorithms to define diabetes and dementia, but we acknowledge that completely accurate diagnoses of dementia in particular, remain a challenge. However, the visual memory test used for follow-up (and thus to define cognitive decline) did not show good reliability (r=0.16) in UKB. UKB had a low response rate and as a result, may suffer from selection bias, which could mean participants were less likely to have cognitive problems at study inception. Thus, it is possible that the association between glycaemia and our outcomes may have been underestimated.

In conclusion, we show that both pre-diabetes and known diabetes are detrimental in terms of vascular dementia risk and cognitive decline and these excess risks appear to be driven by treated hypertension. Somewhat weaker associations with AD and hippocampal volumes indicate pathological mechanisms beyond standard CVD risk factors, in association with hyperglycaemia, that affect brain health. Our findings of potential beneficial effects of low-normal HbA_1c_ on white matter and hippocampal volume are intriguing and require further investigation.

## Data Availability

Data to support this study are available directly from UK Biobank.

## ACKNOWLEGDEMENTS

This work was conducted under the approved UK Biobank project number 7661. We thank the volunteer participants of the UK Biobank, and the UK Biobank researchers.

## AUTHOR CONTRIBUTIONS

Literature search: VG; study design: VG, NC; data analysis: VG, SVE, A-EF; data interpretation: VG, NC, LS, KB; Writing: VG, NC; commenting on the draft: VG, A-EF, SVE, RM, CTR, KB, LS, NC. VG guarantees the work carried out, had access to all of the data and takes responsibility for the integrity of the data and the accuracy of the data analysis. The UK Biobank data are publicly available to all bona fide researchers at https://www.ukbiobank.ac.uk.

## DUALITY OF INTEREST

KB reports grants from Diabetes UK, grants from British Heart Foundation, during the conduct of the study; grants from Medical Research Council, outside the submitted work. LS reports grants from BHF and Diabetes UK, during the conduct of the study; grants from Wellcome, grants from MRC, grants from NIHR, grants from GSK, grants from BHF, outside the submitted work; and is a Trustee of the British Heart Foundation. NC reports grants from Diabetes UK, grants from British Heart Foundation, during the conduct of the study; personal fees from AstraZeneca, grants from the Medical Research Council, outside the submitted work. The remaining authors declare that there are no conflicts of interest.

## FUNDING

This work was jointly funded by Diabetes UK and British Heart Foundation grant 15/0005250. KB holds a Sir Henry Dale Fellowship funded by Wellcome and the Royal Society (grant number 107731/Z/15/Z).

## REFERENCES

1. Rory J McCrimmon, Christopher M Ryan BMF. Diabetes and Cognitive Dysfunction. Lancet 2012;379:2291–99.

2. Xue M, Xu W, Ou YN, et al. Diabetes mellitus and risks of cognitive impairment and dementia: A systematic review and meta-analysis of 144 prospective studies. Ageing Res. Rev. 2019;55:100944.

3. Ravona-Springer R, Luo X, Schmeidler J, et al. Diabetes is associated with increased rate of cognitive decline in questionably demented elderly. Dement. Geriatr. Cogn. Disord. 2010;29(1):68–74.

4. Rosenberg J, Lechea N, Pentang GN, Shah NJ. What magnetic resonance imaging reveals – A systematic review of the relationship between type II diabetes and associated brain distortions of structure and cognitive functioning. Front. Neuroendocrinol. 2018;1–34.

5. Lyall DM, Celis-morales CA, Anderson J, et al. Associations between single and multiple cardiometabolic diseases and cognitive abilities in 474 129 UK Biobank participants. Eur. Heart. J. 2017; 38(8):577–583.

6. Ryan CM, Geckle M. Why is learning and memory dysfunction in Type 2 diabetes limited to older adults?. Diabetes. Metab. Res. Rev. 2000;16(5):308– 315.

7. Biessels GJ, Nobili F, Teunissen CE, et al. Understanding multifactorial brain changes in type 2 diabetes: a biomarker perspective. Lancet Neurol. 2020;19(8):699–710.

8. Gold SM, Dziobek I, Sweat V, et al. Hippocampal damage and memory impairments as possible early brain complications of type 2 diabetes. Diabetologia 2007;50(4):711–719.

9. Fotuhi M, Do D, Jack C. Modifiable factors that alter the size of the hippocampus with ageing [Internet]. Nat. Rev. Neurol. 2012;8(4):189–202.

10. Geijselaers SLC, Sep SJS, Stehouwer CDA, Biessels GJ. Glucose regulation, cognition, and brain MRI in type 2 diabetes: A systematic review [Internet]. Lancet Diabetes Endocrinol. 2015;3(1):75–89.

11. Wardlaw JM, Valdé s Hernández MC, Muñoz-Maniega S. What are white matter hyperintensities made of? Relevance to vascular cognitive impairment. J. Am. Heart Assoc. 2015;4(6):001140.

12. Mankovsky B, Zherdova N, van den Berg E, et al. Cognitive functioning and structural brain abnormalities in people with Type 2 diabetes mellitus. Diabet. Med. 2018;35(12):1663–1670.

13. Sudlow C, Gallacher J, Allen N, et al. UK Biobank: An Open Access Resource for Identifying the Causes of a Wide Range of Complex Diseases of Middle and Old Age. PLoS Med. 2015;12(3):1–10.

14. Eastwood S V, Mathur R, Atkinson M, et al. Algorithms for the Capture and Adjudication of Prevalent and Incident Diabetes in UK Biobank. 2016;

15. Tierney A, Fry D, Almond R, et al. UK Biobank Biomarker Enhancement Project Companion Document to Accompany HbA1c Biomarker Data. 2018;1–8.

16. Ginde AA, Cagliero E, Nathan DM, Camargo CA. Value of Risk Stratification to Increase the Predictive Validity of HbA1c in Screening for Undiagnosed Diabetes in the US Population. 2008;1346–1353.

17. Wilkinson T, Schnier C, Bush K, et al. Identifying dementia outcomes in UK Biobank□: a validation study of primary care, hospital admissions and mortality data. Eur. J. Epidemiol. 2019; 34:557–565.

18. Alfaro-almagro F, Jenkinson M, Bangerter NK, et al. Image processing and Quality Control for the first 10, 000 brain imaging datasets from UK Biobank. NeuroImage 2018;166:400–424.

19. Lyall DM, Cullen B, Allerhand M, et al. Cognitive test scores in UK biobank: Data reduction in 480,416 participants and longitudinal stability in 20,346 participants. PLoS One 2016;11(4):1–10.

20. Frerichs RJ, Tuokko HA. A comparison of methods for measuring cognitive change in older adults. Arch. Clin. Neuropsychol. 2005;20(3):321–333.

21. International Standard Classification of EducationI S C E D 1997.

22. Xue M, Xu W, Ou YN, et al. Diabetes mellitus and risks of cognitive impairment and dementia: A systematic review and meta-analysis of 144 prospective studies. Ageing Res. Rev. 2019;55(January):100944.

23. Euser SM, Sattar N, Witteman JCM, et al. A prospective analysis of elevated fasting glucose levels and cognitive function in older people: Results from PROSPER and the Rotterdam Study. Diabetes 2010;59(7):1601–1607.

24. Marseglia A, Fratiglioni L, Kalpouzos G, et al. Prediabetes and diabetes accelerate cognitive decline and predict microvascular lesions: A population-based cohort study. Alzheimer’s Dement. 2019;15(1):25–33.

25. Glumer C, Jorgensen T, Borch-Johnsen K. Prevalences of diabetes and impaired glucose regulation in a Danish population: the Inter99 study. Diabetes Care 2003;26(8):2335–2340.

26. Holman RR, Paul SK, Bethel MA, et al. 10-year follow-up of intensive glucose control in type 2 diabetes. N. Engl. J. Med. 2008;359(15):1577–1589.

27. Qiu C, Winblad B, Fratiglioni L. T he age-dependent relation of blood pressure to cognitive function and dementia [Internet]. Lancet Neurol. 2005;4(8):487– 499.

28. Whitmer RA, Sidney S, Selby J, et al. Midlife cardiovascular risk factors and risk of dementia in late life. Neurology 2005;64(2):277 LP–281.

29. Lane CA, Barnes J, Nicholas JM, et al. Associations between Vascular Risk across Adulthood and Brain Pathology in Late Life: Evidence from a British Birth Cohort. JAMA Neurol. 2019;1–9.

30. Van Middelaar T, Argillander TE, Schreuder FHBM, et al. Effect of antihypertensive medication on cerebral small vessel disease: A systematic review and meta-analysis. Stroke 2018;49(6):1531–1533.

31. Yano Y, Reis JP, Lewis CE, et al. Association of Blood Pressure Patterns in Young Adulthood With Cardiovascular Disease and Mortality in Middle Age. JAMA Cardiol. 2020; 5(4):382–389.

32. Lane CA, Barnes J, Nicholas JM, et al. Associations between blood pressure across adulthood and late-life brain structure and pathology in the neuroscience substudy of the 1946 British birth cohort (Insight 46): an epidemiological study. Lancet Neurol. 2019;18(10):942–952.

33. Doney ASF, Bonney W, Jefferson E, et al. Investigating the relationship between type 2 diabetes and dementia using electronic medical records in the GoDARTS bioresource. Diabetes Care 2019;42(10):1973–1980.

34. Kisler K, Nelson AR, Montagne A, Zlokovic B V. Cerebral blood flow regulation and neurovascular dysfunction in Alzheimer disease. Nat. Rev. Neurosci. 2017;18(7):419–434.

35. Subramanian N, Chandrasekar S. REACTION TIME IN CLINICAL DIABETES MELLITUS. 1984;2–5.

36. Hainer V, Aldhoon-Hainerová I. Obesity paradox does exist. Diabetes Care 2013;36(SUPPL.2)

